# Multi-Platform Comparison of Plasma Phosphorylated Tau Assays: Analytical and Workflow Factors Drive Clinical Implementation Decisions

**DOI:** 10.64898/2026.05.04.26352262

**Authors:** Olga Sandoval-Boczkowska, John R. Best, Rachael J. Y. Smith, Laura Burns, Cyril Helbling, Ging-Yuek Robin Hsiung, Jacqueline A. Pettersen, Philip E. Lee, Alexandre Henri-Bhargava, Haakon B. Nygaard, Mari L. DeMarco

## Abstract

Multiple plasma phosphorylated tau assays are now commercially available for detection of Alzheimer’s disease (AD) pathology, yet clinical laboratories lack a comprehensive comparative evaluation to guide implementation decisions. Diagnostic accuracy and analytical performance were assessed in a cohort of 273 participants with paired EDTA plasma and CSF specimens. CSF AD core biomarkers were used as the reference standard, and index tests included three plasma pTau217 assays by Roche, Fujirebio, and Meso Scale Discovery [MSD], and a pTau181 assay by Roche. Participants had a median age of 70 [IQR: 64-76] years, 42% were female and 60% were AD-positive. Diagnostic performance was statistically similar across all pTau217 assays (range: 0.88-0.89 area under the receiver operating characteristic curve [AUC]) with the pTau181 assay having lower accuracy (AUC = 0.85). All assays were resistant to hemolysis, icterus, and lipemia. Automated assays (Roche, Fujirebio) showed superior analytical precision and freeze/thaw stability (≥6 cycles) compared to the manual MSD assay (2 cycles). Given that plasma pTau217 assays demonstrated high and comparable accuracy in this head-to-head comparison, their differences in analytical performance characteristics and general clinical laboratory suitability became the differentiating factors for clinical implementation.

## 1 Introduction

Blood biomarkers for Alzheimer’s disease (AD) have emerged with sufficient diagnostic performance to be incorporated into the workup of patients with cognitive impairment suspected to be due to AD, offering a less invasive and more scalable alternative to amyloid positron emission tomography (PET) and cerebrospinal fluid (CSF) analysis, the current gold-standard ante-mortem approaches.[1] Among these, specific proteoforms of tau phosphorylated at residue 217 (pTau217) and 181 (pTau181) have demonstrated particular promise, with pTau217—alone or in combination with other biomarkers—consistently demonstrating the highest diagnostic accuracy [2, 3].

Clinical implementation, however, requires more than strong diagnostic performance. Medically accredited laboratories must also verify analytical performance and operational suitability — yet these considerations are rarely addressed comprehensively in diagnostic accuracy studies, leaving a practical gap for implementing laboratories. Furthermore, medical decision limits must be either verified, if the product has regulatory approval, or established *de novo* in a cohort reflective of the intended use population.

With this in mind, we undertook a head-to-head evaluation of three plasma pTau217 products (Fujirebio, Roche, and Meso Scale Discovery [MSD]) and one pTau181 product (Roche), selected based on reported diagnostic accuracy, availability, regulatory status, and suitability for use within medical laboratories. Beyond diagnostic performance, we characterized analytical performance and operational workflows, and established medical decision limits for all assays using a two-cutpoint system tuned to 90% sensitivity and 90% specificity for detection of AD pathology.

## 2 Methods

### 2.1 Study Design

This study was conducted as an analytical validation and diagnostic accuracy evaluation of commercially available plasma pTau assays, with consideration for implementation in the Canadian health care system. Analytical validation followed Clinical and Laboratory Standards Institute (CLSI) guidelines, and diagnostic accuracy reporting adhered to Standards for Reporting of Diagnostic Accuracy Studies (STARD) criteria [4]. This study was approved by the University of British Columbia and Providence Health Care Research Institute ethics review board.

Plasma pTau assays were selected based on availability within Canada at the time of this study and general suitability for future clinical use. Criteria for clinical suitability included: availability of reagents from established *in vitro* diagnostic (IVD) manufacturers; use of platforms already operating IVD assays in clinical laboratories; and, for research-use-only (RUO) manufacturers, a demonstrated history of instrument use by clinical laboratories as laboratory-developed tests (LDTs). At the time of this study there were no regulatory approved plasma pTau assays available in Canada—this remains true at the time of preparation of this publication. Based on these criteria, assays and instruments evaluated included:

- Roche Elecsys Phospho-Tau (217P) Plasma RUO analyzed on a cobas e402
- Fujirebio Lumipulse G pTau217 analyzed on the Lumipulse G1200
- Meso Scale Discovery (MSD) S-PLEX Human Tau (pT217) analyzed on a MESO SECTOR S600 with data processed using Discovery Workbench, and,
- Roche Elecsys Phospho-Tau (181P) Plasma RUO analyzed on a cobas e402.

### 2.2 Diagnostic accuracy and medical decision limits

All participants, or their substitute decision maker, provided informed consent to participate. Participant inclusion criteria were: availability of an EDTA plasma specimen and paired CSF AD biomarker data. Exclusion criteria were: insufficient plasma volume for testing on all four assays, and/or visual hemolysate exceeding the equivalent of ~0.5 g/L of hemoglobin [5]. It was prespecified that samples exceeding assay-specific acceptability thresholds (determined following evaluation of all common matrix interferents as part of the analytical validation), would be excluded during data analysis.

Specimens and associated data were from one of the following two cohorts: the Canadian Consortium on Neurodegeneration in Ageing’s Comprehensive Assessment of Neurodegeneration and Dementia (COMPASS-ND) study and the Clinic for Alzheimer’s Disease and Related Disorders (CARD) in Vancouver, Canada. COMPASS-ND is a longitudinal, pan-Canadian research cohort of persons aged 50 to 90 spanning both the cognitive spectrum—from cognitively unimpaired (CU) to dementia—and the spectrum of neurodegenerative disorders [6, 7]. The study had 32 enrollment sites across Canada (15 contributing to samples in this study) and set recruitment targets by disease severity, suspected pathology and sex. Participants underwent standardized phenotyping, including neuropsychological testing, functional assessments, neuroimaging, as well as biofluid collection. Recruitment for the CARD study occurred from a single specialized memory clinic that serves as the major neurodegenerative disease referral center in the Canadian province of British Columbia (BC). The clinic is referred patients with suspected or established neurodegenerative disorders. Specimen collection, processing and storage for both cohorts are found in the **Supplementary Data**. Given the respective recruitment catchments for these studies, we refer to them herein as simply the ‘Canada’ and ‘BC’ cohorts.

All participants were classified as ‘positive’ and ‘negative’ for AD pathology based on CSF AD core biomarkers (reference standard). CSF was collected into polypropylene tubes, and aliquoted (500 µL) into polypropylene cryovials for storage at −70 °C prior to analysis (see **Supplementary Data** for details). Samples had been previously analyzed for CSF AD core biomarkers and for reference classification, platform-specific cut-offs were applied. Positivity for AD pathology was defined as follows: Fujirebio Innotest pTau181/Aβ42 >0.08 and/or tTau/Aβ42 >0.52 [8]; Fujirebio Lumipulse Aβ42/Aβ40 <0.063 [9]; LC–MS/MS (Aβ42, Aβ40) and Euroimmun (tTau): Aβ42/Aβ40 <0.071, and/or tTau/Aβ42 >0.28 [10–12]; Roche Elecsys (generation 1) pTau181/Aβ42 >0.024 and/or tTau/Aβ42 >0.29 [13]; Roche Elecsys (generation 2) pTau181/Aβ42 >0.023 and/or tTau/Aβ42 >0.28 [12].

All plasma analyses were performed following the manufacturer’s instructions except for the MSD assay where testing was performed in singleton (versus the recommended duplicate) to enable a fair comparison between platforms. Immediately prior to analysis, plasma aliquots were thawed at room temperature. For the accuracy study, all aliquots underwent a maximum of two freeze–thaw cycles prior to analysis.

### 2.3 Analytical validation: precision, freeze/thaw stability and matrix interference

Following CLSI guideline EP15 [14], precision was assessed using a 5×5 design (five replicates per run across five days, totaling 25 measurements per analyte). Pooled EDTA plasma was aliquoted into 1 mL polypropylene cryovials and stored at −70 °C until use. Within-run and between-run precision were reported as means, standard deviations, and coefficient of variation (CV%).

Following CLSI guideline EP25 [15], analyte stability for each assay was evaluated up to 6 freeze-thaw cycles with percent recovery calculated as the ratio to the baseline concentration. All analyses were performed in duplicate using a common EDTA plasma pool.

Following CLSI guideline C56 [5], spike and recovery experiments were performed to assess interference from hemolyzed, lipemic and icteric specimens. All analyses were performed in duplicate, where 1900 µL of a common EDTA plasma pool was spiked with 100 µL of varying mixtures of phosphate-buffered saline (PBS) and the interferent (i.e., hemolysate from lysed and washed red blood cells, intralipid, bilirubin). Final sample sets had the following interferent concentrations, in addition to the blank spike: 0.50, 1.50, 2.50, and 5.25 g/L of hemoglobin (estimated based on visual inspection [5]); 29.1, 112.9, 273.6, and 513.0 µmol/L of bilirubin, and 125, 250, 500, and 1000 mg/dL of intralipid. Percent recovery was calculated relative to the control containing a blank spike.

### 2.4 Data analysis

Diagnostic accuracy and medical decision limit calculations were prespecified primary analyses. Subgroup analyses stratified by age, sex, education, cognitive stage, APOE-ε4 carrier status, and eGFR were conducted as exploratory analyses to inform future studies and should be interpreted accordingly. Data processing and statistical modelling were performed using R version 4.5.2. Data were descriptively summarized as numbers (and percentages) for categorical variables and as medians (and interquartile ranges) for continuous measures.

Receiver-operating characteristic (ROC) analyses and the computation of area under the ROC curve with bootstrapping (20,000 bootstraps) tested biomarker accuracy. Bootstrapping (20,000 bootstraps) was used to conduct pairwise comparisons of ROC curves for each of the biomarkers and to calculate 95% confidence intervals for the percentage of the study sample within the intermediate range for a given biomarker test. Medical decision limits for the plasma assays were determined using a two cut-point model, in which lower and upper thresholds were selected to achieve ≥90% sensitivity and ≥90% specificity. Non-parametric tests (Wilcoxon rank sum test; Pearson’s chi-squared test, Fisher’s exact test) were used to compare sample characteristics across the two cohorts and by demographic and clinical variables.

### 2.5 Materials

Equipment used included a cobas e402 (Roche Diagnostics, Germany), a Lumipulse G1200 (Fujirebio Inc., Japan), and the MESO SECTOR S600 with Discovery Workbench software (Meso Scale Discovery, USA). Reagents from Roche Diagnostics included the Elecsys Phospho-Tau (217P) (#10285583430) and Phospho-Tau (181P) (#09697870190) plasma assays with associated controls and calibrators. Reagents from Fujirebio included Lumipulse G pTau217 Plasma Immunoreaction Cartridges (#81472) with associated controls and calibrators. Reagents from MSD included the S-PLEX Human Tau (pT217) kit (#K151APFS-1). Additional materials used included PBS tablets (#P4417-50TAB; Sigma-Aldrich, USA), unconjugated bilirubin (CEQAL Inc., Canada), intralipid (#831818610, Fresenius Kabi Canada Ltd., Canada) and EDTA plasma pools (anonymized specimens from St. Paul’s Hospital Laboratory, Vancouver, Canada).

## 3 Results

### 3.1 Patient participants

The study cohort comprised 273 participants with a median age of 70 years (IQR: 64–76); 42% were female and 60% were positive for AD pathology, with the majority at the stage of mild cognitive impairment (49%) (**Table 1**). Comparing between the BC and Canada cohorts revealed the following: a higher AD prevalence in the BC cohort (70% v. 53%; *p* = 0.005 for Chi-squared test), marginally higher prevalence of APOE-ε4 carrier status in the BC cohort (50% vs. 44%; p = 0.047 for difference), and no statistical difference in median pTau217 or pTau181 concentrations between cohorts (*p* > 0.30).

**Table 1.**
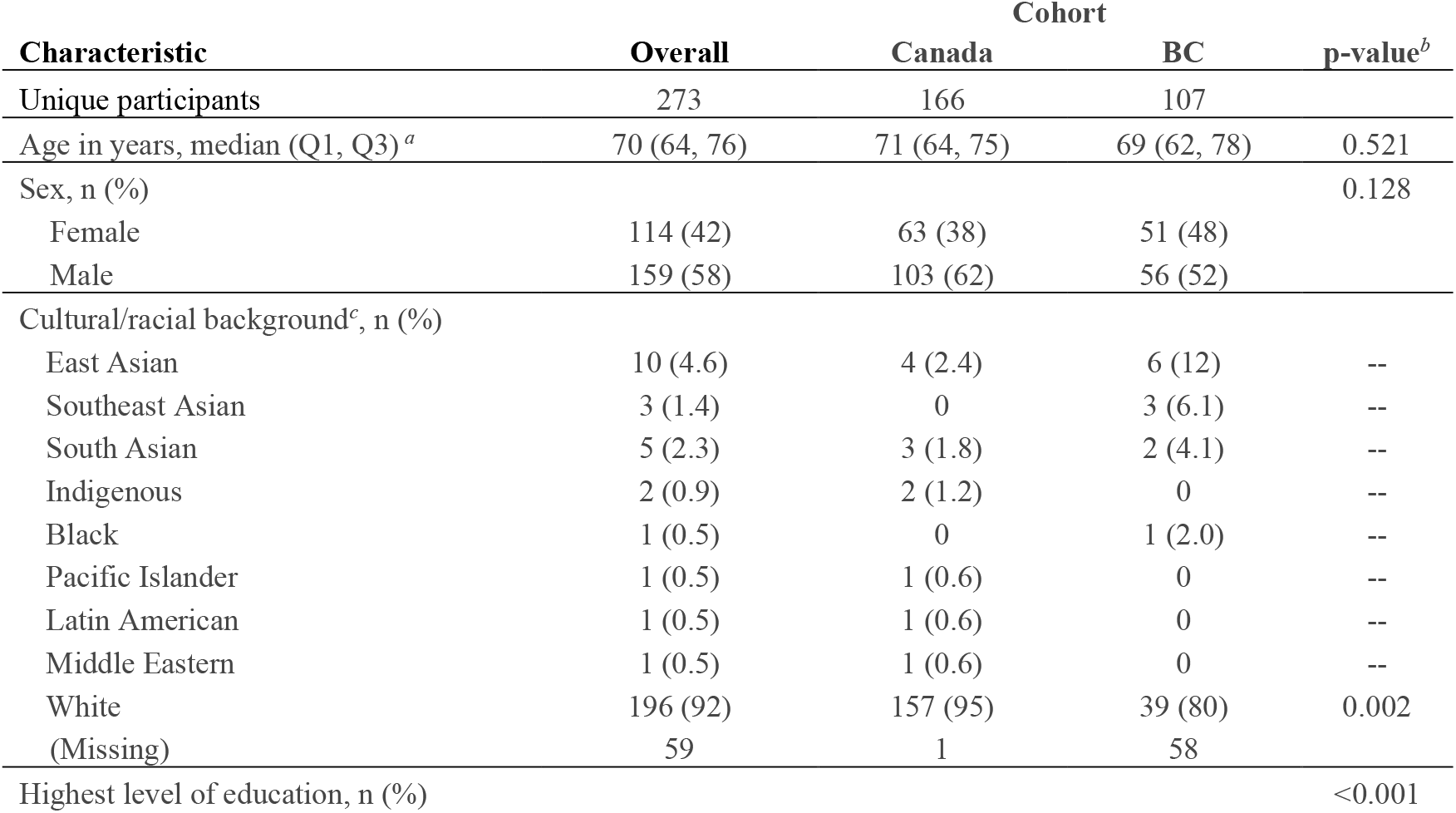
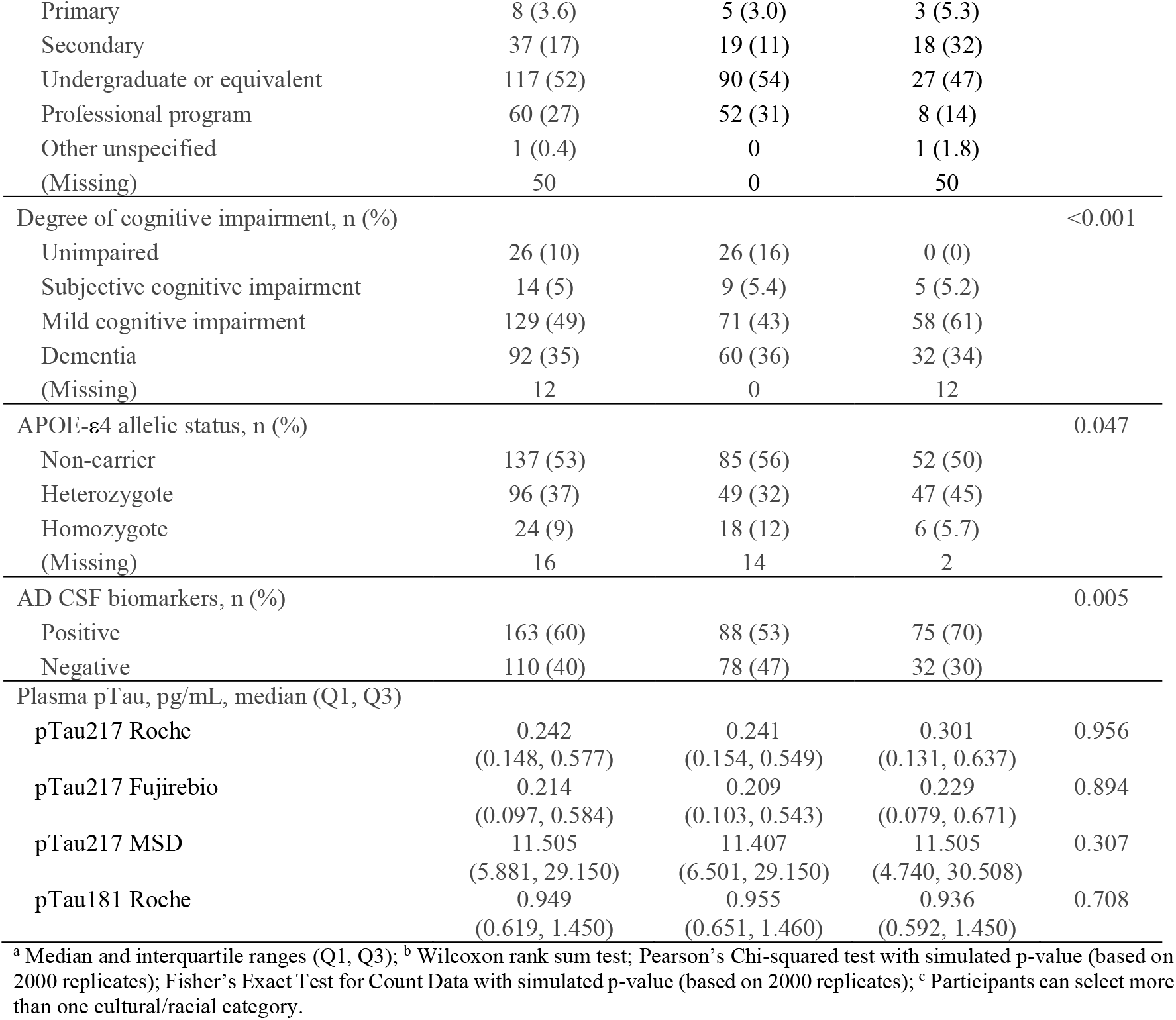
Demographic and biomarker data of included participants.

| Characteristic | Overall | Cohort |  | p-value<sup>b</sup> |
| --- | --- | --- | --- | --- |
|  |  | Canada | BC |  |
| Unique participants | 273 | 166 | 107 |  |
| Age in years, median (Q1, Q3)<sup>a</sup> | 70 (64, 76) | 71 (64, 75) | 69 (62, 78) | 0.521 |
| Sex, n (%) |  |  |  | 0.128 |
| Female | 114 (42) | 63 (38) | 51 (48) |  |
| Male | 159 (58) | 103 (62) | 56 (52) |  |
| Cultural/racial background<sup>c</sup>, n (%) |  |  |  |  |
| East Asian | 10 (4.6) | 4 (2.4) | 6 (12) | -- |
| Southeast Asian | 3 (1.4) | 0 | 3 (6.1) | -- |
| South Asian | 5 (2.3) | 3 (1.8) | 2 (4.1) | -- |
| Indigenous | 2 (0.9) | 2 (1.2) | 0 | -- |
| Black | 1 (0.5) | 0 | 1 (2.0) | -- |
| Pacific Islander | 1 (0.5) | 1 (0.6) | 0 | -- |
| Latin American | 1 (0.5) | 1 (0.6) | 0 | -- |
| Middle Eastern | 1 (0.5) | 1 (0.6) | 0 | -- |
| White | 196 (92) | 157 (95) | 39 (80) | 0.002 |
| (Missing) | 59 | 1 | 58 |  |
| Highest level of education, n (%) |  |  |  | <0.001 |
| Primary | 8 (3.6) | 5 (3.0) | 3 (5.3) |  |
| Secondary | 37 (17) | 19 (11) | 18 (32) |  |
| Undergraduate or equivalent | 117 (52) | 90 (54) | 27 (47) |  |
| Professional program | 60 (27) | 52 (31) | 8 (14) |  |
| Other unspecified | 1 (0.4) | 0 | 1 (1.8) |  |
| (Missing) | 50 | 0 | 50 |  |
| Degree of cognitive impairment, n (%) |  |  |  | <0.001 |
| Unimpaired | 26 (10) | 26 (16) | 0 (0) |  |
| Subjective cognitive impairment | 14 (5) | 9 (5.4) | 5 (5.2) |  |
| Mild cognitive impairment | 129 (49) | 71 (43) | 58 (61) |  |
| Dementia | 92 (35) | 60 (36) | 32 (34) |  |
| (Missing) | 12 | 0 | 12 |  |
| APOE-ε4 allelic status, n (%) |  |  |  | 0.047 |
| Non-carrier | 137 (53) | 85 (56) | 52 (50) |  |
| Heterozygote | 96 (37) | 49 (32) | 47 (45) |  |
| Homozygote | 24 (9) | 18 (12) | 6 (5.7) |  |
| (Missing) | 16 | 14 | 2 |  |
| AD CSF biomarkers, n (%) |  |  |  | 0.005 |
| Positive | 163 (60) | 88 (53) | 75 (70) |  |
| Negative | 110 (40) | 78 (47) | 32 (30) |  |
| Plasma pTau, pg/mL, median (Q1, Q3) |  |  |  |  |
| pTau217 Roche | 0.242<br>(0.148, 0.577) | 0.241<br>(0.154, 0.549) | 0.301<br>(0.131, 0.637) | 0.956 |
| pTau217 Fujirebio | 0.214<br>(0.097, 0.584) | 0.209<br>(0.103, 0.543) | 0.229<br>(0.079, 0.671) | 0.894 |
| pTau217 MSD | 11.505<br>(5.881, 29.150) | 11.407<br>(6.501, 29.150) | 11.505<br>(4.740, 30.508) | 0.307 |
| pTau181 Roche | 0.949<br>(0.619, 1.450) | 0.955<br>(0.651, 1.460) | 0.936<br>(0.592, 1.450) | 0.708 |

<sup>a</sup> Median and interquartile ranges (Q1, Q3); <sup>b</sup> Wilcoxon rank sum test; Pearson's Chi-squared test with simulated p-value (based on 2000 replicates); Fisher's Exact Test for Count Data with simulated p-value (based on 2000 replicates); <sup>c</sup> Participants can select more than one cultural/racial category.

### 3.2 Diagnostic accuracy and medical decision limits

All assays demonstrated high discriminative ability for differentiating individuals with AD pathology (**Figure 1**). AUCs for pTau217 were all similar, ranging from 0.88 to 0.89 with overlapping confidence intervals (**Figure 1**). The pTau181 assay demonstrated slightly lower discriminative ability (AUC: 0.85, **Table S2**). Absolute AUC differences (ΔAUC) from the bootstrap test for two correlated ROC curves ranged from 0.024 to 0.033, with corresponding absolute test statistics (|D|) 1.58–2.01 and p-values 0.044–0.11 (**Table S1**). The smallest p-values were observed when comparing Roche pTau181 to Fujirebio pTau217 (ΔAUC=0.033, |D|=2.01, p=0.044) and to Roche pTau217 (ΔAUC=0.026, |D|=1.86, p=0.063). All other pairwise comparisons among pTau217 assays yielded ΔAUC=0.003–0.010 with p≥0.48.

**Figure 1.**
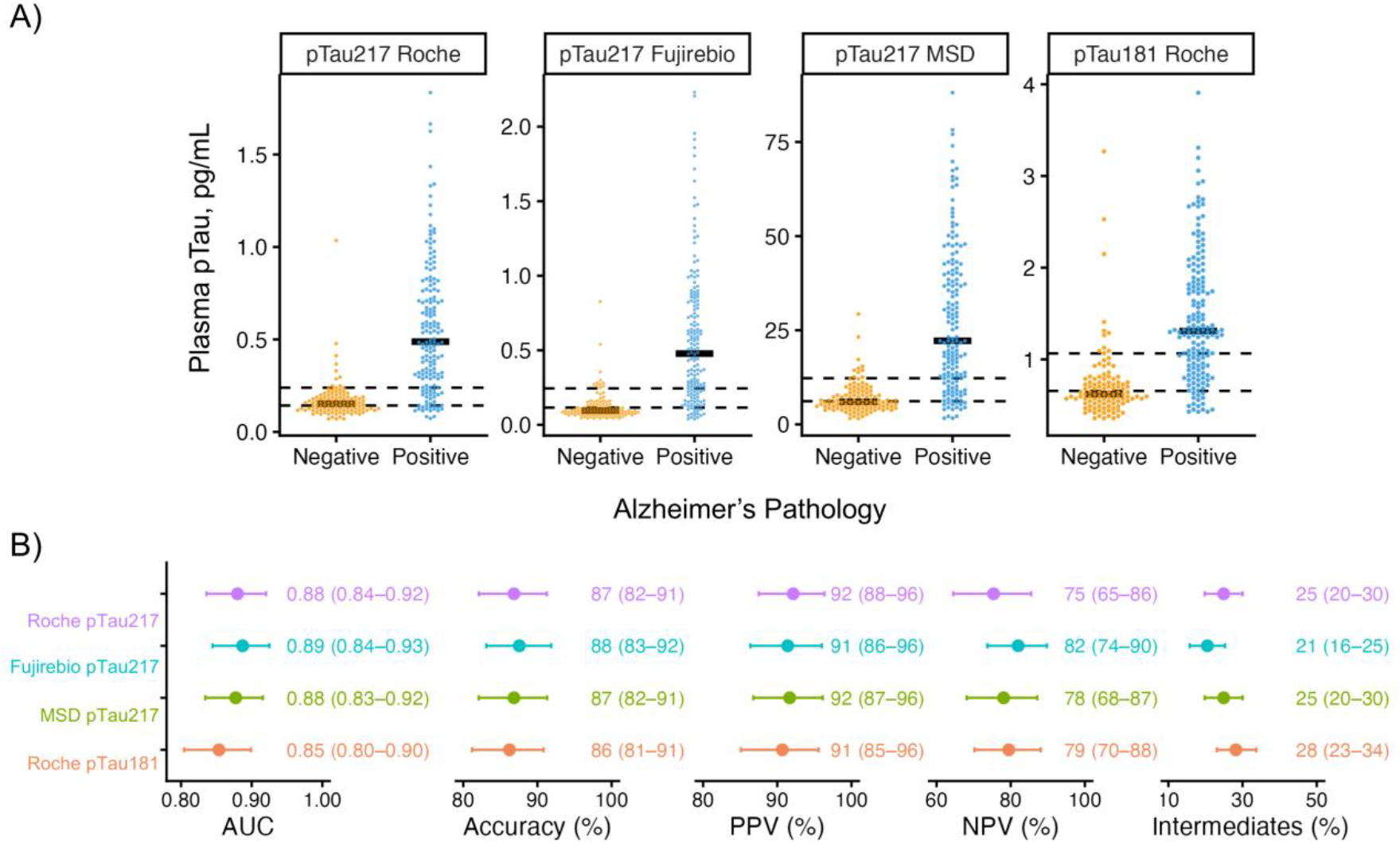
Comparative performance of plasma phosphorylated tau (pTau) 217 and 181 assays in a large Canadian cohort (n=273) with medical decision limits defined at 90% sensitivity and 90% specificity for detection of Alzheimer’s pathology. (A) Distribution of plasma pTau results by AD pathology status (yellow: negative; blue: positive), with medical decision limits (dotted lines) and median values (solid lines) shown. (B) Area under the receiver operating characteristic curve (AUC), accuracy, positive predictive value (PPV), negative predictive value (NPV), and intermediate cases for each pTau assay, with 95% confidence intervals.

Medical decision limits were calculated *de novo* for each assay, with all calculations including all 273 participant pTau results (**Figure 1A & Table S2**). All assays demonstrated high inter-assay concordance with ρ ranging from 0.893 (Roche pTau181 v. MSD pTau217) to 0.925 (Roche v. MSD pTau217) (**Figure S1**).

### 3.3 Exploratory analyses

Performance by cohort (**Table S3**) and by demographic and clinical characteristics was generally consistent with the overall cohort and the relative ordering of assays was preserved across all subgroups (**Figure 2**). No significant AUC differences were observed by cohort (**Table S3**). AUC and accuracy differences were largest for APOE-ε4 carrier status and cognitive stage. AUCs were higher in females than males across all assays, corresponding to a higher AD prevalence in females (67% vs. 55%, *p* = 0.065 for Chi-squared test) and higher pTau concentrations in females (**Table S4**). Age and education subgroups showed modest, overlapping differences. The eGFR ≥90 subgroup (n=44) had lower AUCs than the eGFR 60–89 subgroup (**Figure 2**).

**Figure 2.**
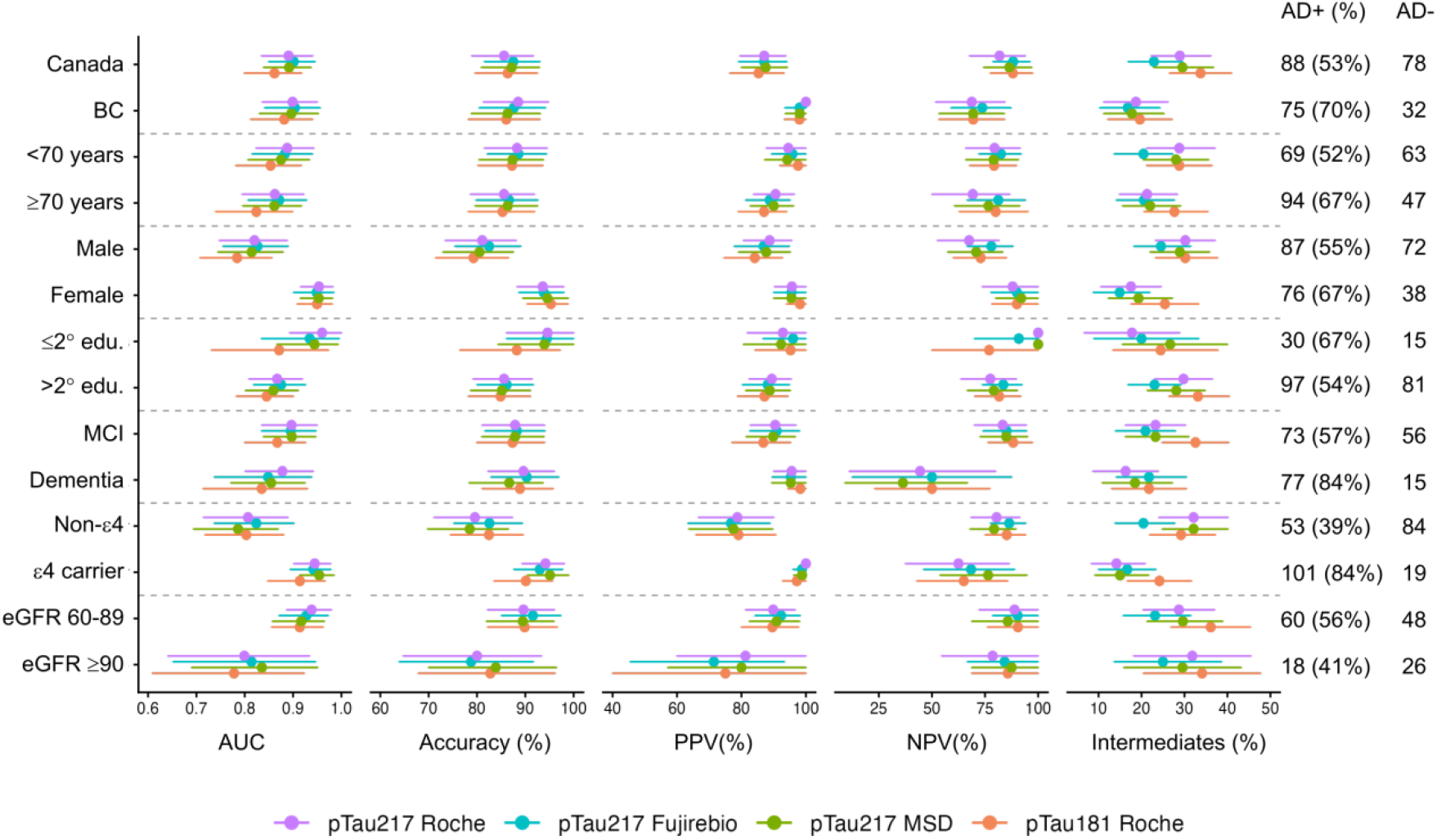
Performance of pTau biomarkers stratified by: age (<70 v. ≥70 years), sex (male v. female), education (≤secondary v. >secondary education), cognitive impairment (mild cognitive impairment [MCI] v. dementia), *APOE*-ε4 carrier status, and estimated glomerular filtration rate (eGFR, mL/min/1.73m^2^). Number of cases with (AD+) and without (AD-) Alzheimer’s disease pathology per subgroup shown, along with percent prevalence of AD+ in each subgroup.

### 3.4 Analytical performance

The automated platforms (Roche and Fujirebio) outperformed the manual platform (MSD) on all analytical performance metrics, with total imprecision ranging from 2.0% to 11.5% for automated assays versus 17.9% for MSD. Results of within-run, between-run and total imprecision testing for all assays are found in **Figure 3**. Performance (percent recovery) of each analyte/assay after freeze/thaw cycles, and in the presence of hemolysate, intralipid and bilirubin is found in **Figure 4**.

**Figure 3.**
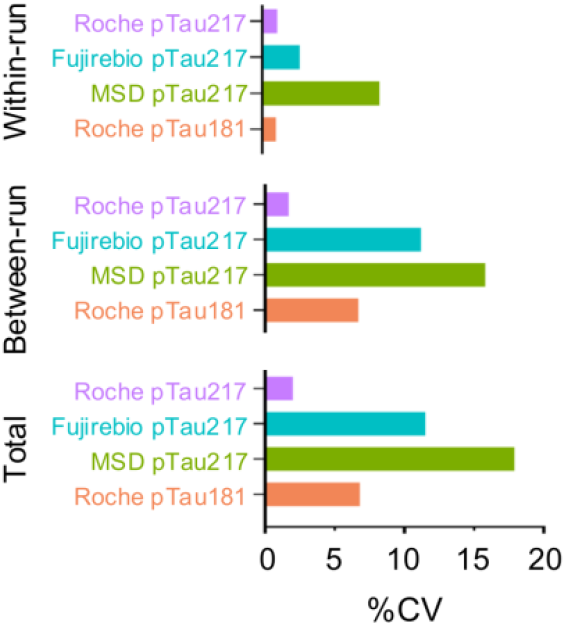
Analytical imprecision from an EDTA plasma pool analyzed across all pTau assays in a 5×5 experiment. Imprecision expressed as within-run coefficient of variation (CV), between-run CV and total CV.

**Figure 4.**
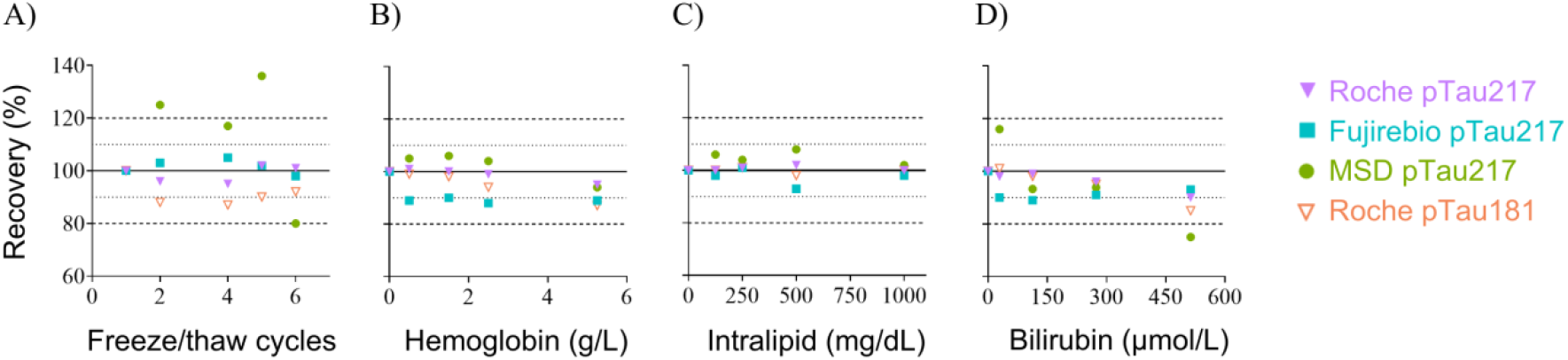
Analyte recovery under a variety of conditions. (A) Automated pTau217 assays remained within ±5% of baseline after up to 6 freeze/thaw cycles, with pTau181 remaining within ±20%, and the one manual assay (MSD pTau217) exceeding 20% after only 2 cycles. (B-D) All assays tested were robust to interference by hemolysate and lipemia, with a trend towards negative interference with icteric specimens.

Instrumentation options, with corresponding assay volume requirements, analytical time, assay throughput, and relative costs are found in **Table 2**.

**Table 2.**
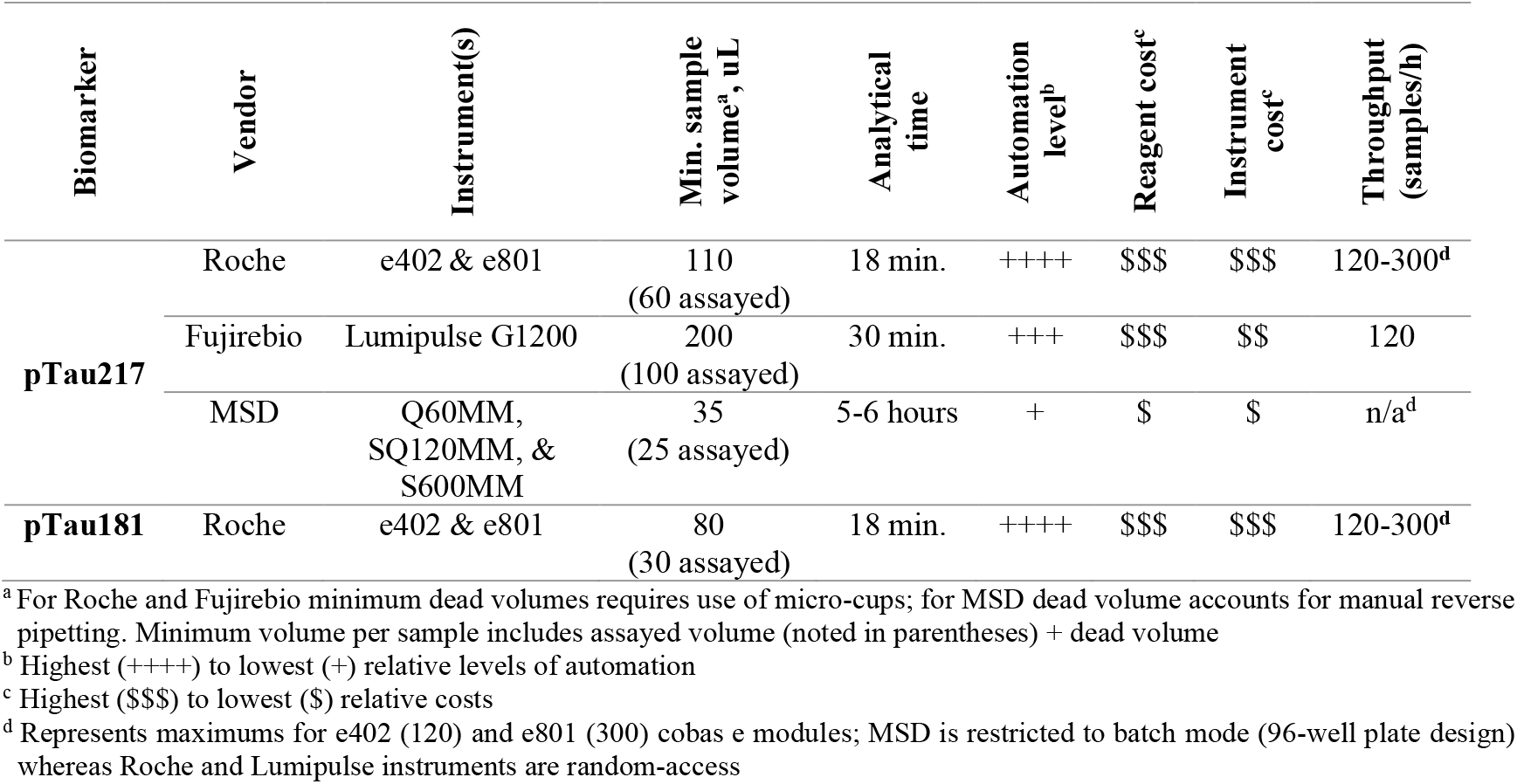
Summary of plasma pTau assay methodology including minimum required plasma volume, analytical time, relative level of automation and costs, and maximum sample throughput per hour.

## 4 Discussion

In a head-to-head comparison of three commercially available plasma pTau217 assays and one pTau181 assay in 273 participants, diagnostic accuracy was statistically comparable across all pTau217 platforms, thus shifting the basis for clinical implementation decisions to analytical performance and operational suitability. Diagnostic accuracy was statistically comparable across all three pTau217 assays (AUC range 0.88-0.89), and pTau181 (AUC 0.85) performed modestly but statistically lower relative to the top performing pTau217 assay. The overall AD pathology prevalence of 60% aligns with intended use of this testing in specialized practice [16]. While the two sub-cohorts differed in AD prevalence, 70% (BC) versus 53% (Canada), AUC were statistically similar across all pTau217 assays. Evaluated against this backdrop of equivalent clinical accuracy, differences in analytical performance and operational workflows emerged as key differentiators for medical laboratories seeking to implement plasma pTau217 testing.

The observation that pTau217 consistently outperforms pTau181 aligns with prior literature [2, 17, 18]. Based on these collective findings, laboratories interested in implementing plasma pTau should prioritize pTau217 over pTau181. AUCs for pTau217 (range: 0.88-0.89) were modestly lower than those reported in large comparative studies of 998 and 1,767 participants.[19, 20] Differences in AUC likely reflect cohort composition and reference standard differences, rather than true assay performance discrepancies. The first study used amyloid-PET as the reference standard and included a large proportion of cognitively unimpaired individuals (which is likely to improve separation between disease and controls and increase AUC) [19]. The second study included a catchment of 4 secondary care sites and a single CSF reference standard whereas our study included a catchment of 16 secondary and tertiary care sites and comparison to multiple CSF methods [20]. A narrower enrollment catchment and single comparator is likely to yield lower variability; whereas a higher number of collection sites and comparator methods better estimate performance across health systems.

In stratifying performance by demographic and clinical variables, in general, we observed the preservation of relative assay ordering supporting the generalizability of the overall performance findings. Subgroup PPV and NPV differences largely tracked with differences in AD prevalence, as expected given the dependence of these metrics on disease prevalence. Subgroup performance evaluation was not a primary aim of this study and would require prospective designs with random sampling. With this caveat in mind, we observed sex differences that mirror previous findings including higher plasma pTau217 in females as well as possible sex-specific differences in the relationship between pTau217 and underlying AD pathophysiology [21], which may contribute beyond the prevalence difference alone. Given previous evidence of the association of chronic kidney disease with increased plasma pTau217 and pTau181 concentrations, we attempted stratification by chronic kidney disease stages. We were only able to stratify by stages 1 (eGFR ≥90; n=44) and 2 (eGFR 60-90; n=108), as there were only 14 participants at or below stage 3. Given the small stage 1 subgroup and subsequent wide confidence intervals, our ability to draw strong conclusions from this analysis was limited. Collectively, these subgroup findings are consistent with prior reports documenting the influence of sex and renal function on plasma pTau217 concentrations [19, 20, 22] and support the relevance of these variables when interpreting results at the individual patient level.

Having established comparable diagnostic performance among pTau217 assays, analytical characteristics become the decisive differentiating factor for clinical laboratory implementation. The Roche pTau217 assay demonstrated the best overall precision, with a total CV of 2.0%. The Fujirebio pTau217 assay showed excellent within-run precision (CV: 2.7%) but exhibited higher between-run (CV: 11.2%) and total imprecision (CV: 11.5%). It is expected that more frequent calibration would largely mitigate this issue. The MSD pTau217 assay demonstrated the highest total imprecision at 17.9%, consistent with the manual sample preparation and plate-based workflow inherent to this platform. While applicable for research purposes (particularly if run in duplicate to reduce imprecision), high imprecision in a clinical setting reduces confidence in medical decision limits and undermines reliability of test interpretation.

The Roche and Fujirebio pTau217 assays maintained analyte recovery within 5% of baseline through six freeze/thaw cycles, making both assays well-suited to routine clinical laboratory workflows where samples may undergo multiple freeze/thaw cycles during processing, shipping/storage or repeat testing. The MSD assay showed unacceptable bias after only two freeze/thaw cycles. Given the high total imprecision of the assay, this failure is likely attributable in large part to measurement variability; however, this still represents a meaningful constraint for clinical use.

All four assays demonstrated general robustness to interference from hemolysis, lipemia and icterus at clinically relevant concentrations. Analyte recovery remained within approximately ±10% at moderate concentrations of hemoglobin, intralipid and bilirubin. Performance degraded across platforms at the highest hemoglobin and bilirubin concentrations tested; such severely abnormal matrices are unlikely to be frequently encountered in routine practice for this patient population and therefore are not anticipated to have a noticeable impact on specimen rejection rates. All assays were notably resilient to interference from lipemia.

Beyond analytical performance, operational suitability is a central consideration for clinical laboratories. The fully automated platforms offer random-access testing, high throughput (up to 120–300 samples per hour), short analytical turnaround (18–30 minutes), and a high degree of automation, enabling integration into existing laboratory workflows with minimal additional hands-on time. The manual platform, by contrast, operates exclusively in batch mode with a 5–6 hour analytical cycle, requiring significant hands-on time for manual plate preparation, though its primary advantages are substantially lower instrument and reagent costs and a low minimum sample volume (35 µL total). A comprehensive comparison of operational parameters to inform platform selection is provided in **Table 2**.

This study has several notable strengths. The cohort was drawn from over 16 sites spanning multiple provinces and cities across Canada, supporting the generalizability of findings beyond a single center or regional population. The use of a multi-method, multi-platform CSF reference standard more accurately captures inter-health system variability than a single comparator method. The cohort’s prevalence of AD pathology, median age, and distribution of cognitive impairment stages are consistent with guidelines for plasma biomarker use in specialized care settings, supporting the external applicability of the derived medical decision limits, while enriching available plasma pTau data with a Canadian cohort. Critically, the evaluation extends beyond diagnostic accuracy to include CLSI-compliant analytical validation, providing data required by clinical laboratories prior to implementation but that most diagnostic accuracy studies do not report. Comprehensive analytical validation data of this kind can be leveraged by implementing laboratories to support verification rather than *de novo* validation, substantially reducing the barrier to implementation—particularly given that establishment of medical decision limits requires paired reference standard data (amyloid PET or CSF) that most laboratories cannot access.

Several limitations warrant acknowledgement. While the use of multiple CSF platforms and methods for reference classification was intended to capture inter-health system variability, it introduces heterogeneity in reference standard assignment that may influence diagnostic accuracy estimates. As paired CSF data were required for reference classification, specimens were drawn from two research cohorts, which were predominantly White and highly educated and lacked sufficient representation of select subgroups for meaningful subgroup analysis (e.g., advanced renal impairment and SCI). At the time of this work, no IVD-approved plasma pTau products existed in Canada, necessitating use of RUO-designated assays and limiting access to certain products in development.

## Data Availability

The data that support the findings of this study are available from the corresponding author upon reasonable request.

## 6 Acknowledgements

We thank all study participants including patients and their care partners as without their contributions this work would not be possible. All authors are members the Canadian Consortium on Neurodegeneration in Aging (CCNA). The CCNA is supported by a grant from the Canadian Institutes of Health Research with funding from several partners.

## 7 Sources of Funding

This research was funded in part by a research grant from the Ministry of Health (Government of British Columbia) to MLD. In kind support (to institution) was provided in part by MSD (reagents), Roche Diagnostics (reagents), and Fujirebio via Phoenix Airmid (reagents and equipment). CH is supported in part by a Four-Year Doctoral Fellowship from University of British Columbia (#6456).

## 8 Disclosures

**OS, JRB, RS, LB** and **CH** declare no conflicts of interest. Outside of the submitted work, the authors report the following disclosures. **AHB**: Grants/contracts paid to institution from ICG Pharma, Alnylam, AriBio, Novo Nordisk, Cerevel, Anavex, the Canadian Consortium on Neurodegeneration in Aging, and CABHI; consulting fees and honoraria from Eli Lilly and Eisai (payments to professional corporation); volunteer board roles with the Consortium of Canadian Centres for Clinical Cognitive Research and the Canadian Neurological Society. **JAP**: Consulting fees and honoraria from Eisai and Eli Lilly. Volunteer board role with Canadian Colloquium on Dementia and scientific advisory committee for CLEAR. **GRH**: Grant funding from Biogen, Roche, Cassava Sciences, and Eisai; consulting for Biogen, Roche, Novo Nordisk, Eisai, and Eli Lilly; serves as President of the Consortium of Canadian Centres for Clinical Cognitive Research. **PEL**: Consulting for Eli Lilly and Eisai. **HBN**: Consulting for Eisai, and advisory boards for Biogen and Hoffmann-La Roche. **MLD** reports consulting for Canada’s Drug Agency, Siemens, Roche and Eisai, lecturing/educational activity fees from Eli Lilly and Roche, and role as co-chair for the Canadian Society of Clinical Chemists/Association for Diagnostics & Laboratory Medicine Guidance Document on Alzheimer’s Disease Biofluid Biomarkers.

## Notes

### Funding Statement

This study was funded in part by a research grant from the Ministry of Health (Government of British Columbia) to MLD. In kind support (to institution) was provided in part by MSD (reagents), Roche Diagnostics (reagents), and Fujirebio via Phoenix Airmid (reagents and equipment). CH is supported in part by a Four-Year Doctoral Fellowship from University of British Columbia.

### Author Declarations

Ethics committee of the University of British Columbia and Providence Health Care Research Institute gave ethical approval for this work.

