## Supplemental Data for "Multi-Platform Comparison of Plasma Phosphorylated Tau Assays: Analytical and Workflow Factors Drive Clinical Implementation Decisions"

### Supplementary Methods

#### *COMPASS-ND (Canada cohort) protocol*

CSF and blood were collected, processed and stored following CCNA study protocols (CCNA Lab Manual v7, 2018). For plasma, blood was collected into 6 mL K2-EDTA tubes (#02-657-32; BD Diagnostics, USA). Following collection, EDTA tubes were gently inverted (10-12 times); tubes were placed on ice and processed within 30 min. Plasma was isolated by centrifugation (3000 rpm, 15 min, 4 °C) and aliquoted into 500 µL polypropylene cryovials (#3743; Thermo Fisher Scientific, USA).

CSF was collected by lumbar puncture with up to 15 mL of CSF collected per participant. The first 1–2 mL was collected into two sterile hospital tubes for routine clinical analyses (cell count, glucose, protein). The subsequent 12 mL was collected into two 10 mL polypropylene tubes (#62.610.201; Sarstedt, Germany). CSF was obtained either by spontaneous drip or by slow withdrawal via syringe (#300912; Becton Dickinson, USA). Following collection, polypropylene tubes were transported to the local biospecimen processing room, logged in the LORIS system, and processed within the same visit. CSF was aliquoted into 500 µL polypropylene cryovials (#3743; Thermo Fisher Scientific, USA), generating up to 24 aliquots per collection and stored at −70 °C.

#### *CARD (BC cohort) protocol*

Blood samples were collected following procedures modeled after ADNI. Blood was drawn into 6 mL K2-EDTA tubes (BD Vacutainer). Tubes were centrifuged within 30 minutes of collection (1500–2500 *g*, 15 min, 4 °C) and plasma was aliquoted into 2 mL polypropylene cryovials (#72.380.007; Sarstedt, Germany), and stored at −70 °C.

CSF was collected by lumbar puncture, typically 15–20 mL per participant, into a sterile polypropylene low-bind tube (Falcon Conical Centrifuge polypropylene tubes, Corning Life Sciences). Within 4 hours of collection, samples were centrifuged at 2,800 g for 10 min at 4 °C, and supernatant aliquoted into 2 mL polypropylene cryovials (#72.380.007, Sarstedt, Germany), and immediately frozen at −80 °C.

### Supplementary Tables

**Table S1.** Comparison of receiver operating characteristic area under curve (AUC) values.

| Assay 1 | Assay 2 | AUC Difference (absolute value) | Test statistic (\|D\|) | p-value |
|---|---|---|---|---|
| **pTau181 Roche** | pTau217 Roche | 0.026 | 1.86 | 0.063 |
| **pTau181 Roche** | pTau217 MSD | 0.024 | 1.58 | 0.11 |
| **pTau181 Roche** | pTau217 Fujirebio | 0.033 | 2.01 | 0.044 |
| **pTau217 Fujirebio** | pTau217 Roche | 0.007 | 0.48 | 0.63 |
| **pTau217 MSD** | pTau217 Roche | 0.003 | 0.22 | 0.83 |
| **pTau217 MSD** | pTau217 Fujirebio | 0.010 | 0.71 | 0.48 |

**Table S2.** Plasma phosphorylated tau (pTau) medical decision limits (MDL) and corresponding sensitivities (SN) and specificities (SP) for detection of Alzheimer's disease pathology.

| Biomarker | Lower MDL at 90% SN | | Upper MDL at 90% SP | |
|---|---|---|---|---|
| | **pg/mL** | **SP, %** | **pg/mL** | **SN, %** |
| **pTau217 Roche** | 0.142 | 45 | 0.239 | 79 |
| **PTau217 Fujirebio** | 0.116 | 66 | 0.244 | 72 |
| **pTau217 MSD** | 6.160 | 52 | 12.271 | 74 |
| **pTau181 Roche** | 0.656 | 56 | 1.065 | 66 |

**Table S3.** Comparison of receiver operating characteristic area under curve (AUC) values by cohort with 95% confidence intervals (CI) and breakdown of participants with and without Alzheimer's disease pathology (AD+ & AD-, respectively).

| | **Canada Cohort** 88 AD+, 78 AD- | | **BC Cohort** 78 AD+, 37 AD- | | **p-value** |
|---|---|---|---|---|---|
| **Plasma Biomarker** | **AUC** | **95% CI** | **AUC** | **95% CI** | |
| pTau217 Roche | 0.89 | 0.84-0.94 | 0.90 | 0.84-0.95 | 0.84 |
| pTau217 Fujirebio | 0.90 | 0.85-0.95 | 0.90 | 0.84-0.96 | 0.95 |
| pTau217 MSD | 0.89 | 0.84-0.94 | 0.90 | 0.83-0.95 | 0.91 |
| pTau181 Roche | 0.86 | 0.80-0.92 | 0.88 | 0.81-0.94 | 0.65 |

**Table S4.** Descriptive statistics stratified by sex.

| Characteristic | Female | Male | p-value[b] |
|---|---|---|---|
| Unique participants | 114 | 159 | |
| Age in years, median (Q1, Q3)[a] | 68 (63, 75) | 71 (64, 77) | 0.114 |
| Cohort, n (%) | | | 0.128 |
| BC | 51 (45) | 56 (35) | |
| Canada | 63 (55) | 103 (65) | |
| Cultural/racial background[c], n (%) | | | |
| East Asian | 7 (8.2) | 3 (2.3) | -- |
| Southeast Asian | 2 (2.4) | 1 (0.8) | -- |
| South Asian | 2 (2.4) | 3 (2.3) | -- |
| Indigenous | 0 | 2 (1.6) | -- |
| Black | 0 | 1 (0.8) | -- |
| Pacific Islander | 0 | 1 (0.8) | -- |
| Latin American | 0 | 1 (0.8) | -- |
| Middle Eastern | 0 | 1 (0.8) | -- |
| White | 76 (89) | 120 (93) | 0.446 |
| (Missing) | 30 | 30 | |
| Highest level of education, n (%) | | | 0.095 |
| Primary | 3 (3.3) | 5 (3.8) | |
| Secondary | 22 (24) | 15 (11) | |
| Undergraduate or equivalent | 44 (48) | 73 (56) | |
| Professional program | 22 (24) | 38 (29) | |
| Other unspecified | 1 (1.1) | 0 | |
| (Missing) | 22 | 28 | |
| Degree of cognitive impairment, n (%) | | | 0.906 |
| Unimpaired | 10 (9.3) | 26 (10) | |
| Subjective cognitive impairment | 7 (6.5) | 7 (4.5) | |
| Mild cognitive impairment | 52 (49) | 77 (60) | |
| Dementia | 33 (36) | 54 (35) | |
| (Missing) | 7 | 5 | |
| APOE-ε4 allelic status, n (%) | | | 0.347 |
| Non-carrier | 53 (49) | 85 (57) | |
| Heterozygote | 45 (41) | 51 (34) | |
| Homozygote | 11 (10) | 13 (8.7) | |
| (Missing) | 6 | 10 | |
| AD CSF biomarkers, n (%) | | | 0.065 |
| Positive | 76 (67) | 87 (55) | |
| Negative | 38 (33) | 72 (45) | |
| Plasma pTau, pg/mL, median (Q1, Q3) | | | |
| pTau217 Roche | 0.317 (0.153, 0.669) | 0.208 (0.138, 0.492) | 0.032 |
| pTau217 Fujirebio | 0.359 (0.106, 0.725) | 0.156 (0.092, 0.406) | 0.002 |
| pTau217 MSD | 14.496 (6.501, 36.874) | 9.210 (5.446, 21.472) | 0.005 |
| pTau181 Roche | 1.020 (0.639, 1.750) | 0.863 (0.604, 1.340) | 0.166 |

[a] Median and interquartile ranges (Q1, Q3); [b] Wilcoxon rank sum test; Pearson's Chi-squared test with simulated p-value (based on 2000 replicates); Fisher's Exact Test for Count Data with simulated p-value (based on 2000 replicates); [c] Individuals can belong to more than one cultural/racial background category. Due to small cell sizes, a between-cohort statistical comparison was only conducted on the White category.

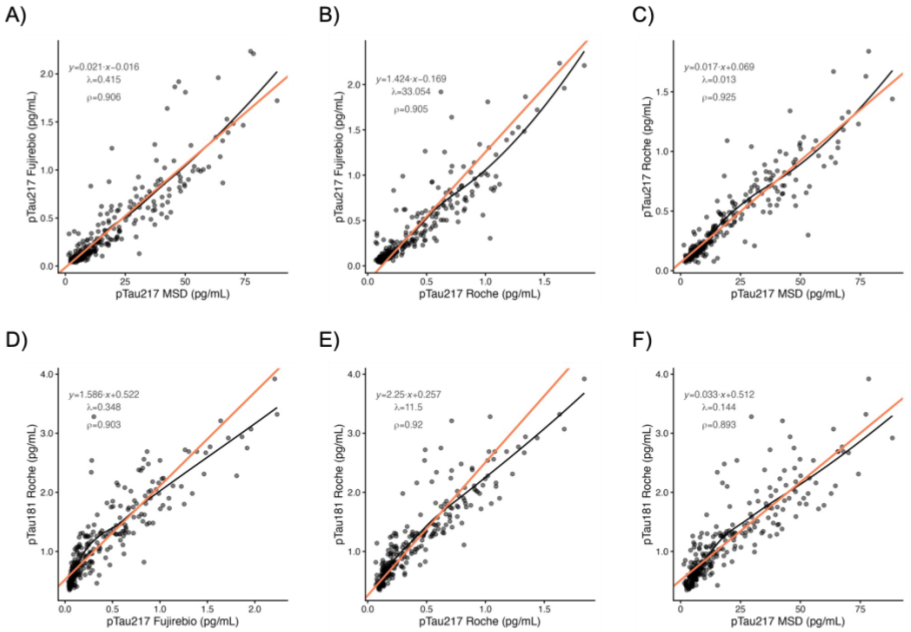


**Figure S1.** Inter-assay concordance. Deming regression (orange line) was used to account for measurement error in both assays, with the error variance ratio (λ) estimated from the precision data. Intercept, slope, and λ values are presented, along with the non-parametric correlation value (ρ) and regression curve (LOESS, black curve).
